# Quantitative interpretation of Sedia LAg Assay test results after HIV diagnosis

**DOI:** 10.1101/2021.09.13.21263495

**Authors:** Joseph Sempa, Eduard Grebe, Alex Welte

## Abstract

**Background:** Testing for ‘recent HIV infection’ is common in surveillance, where only population-level estimates (of incidence) are reported. Typically, ‘recent infection’ is a *category*, obtained by applying a threshold on an underlying continuous biomarker from some laboratory assay(s). Somehow interpreting the biomarker values obtained for individual subjects, for example interpreting them as estimates of the date of infection, has obvious potential applications in the context of studies of early infection, and has also for some years attracted significant interest as an extra component of post-test counselling and treatment initiation. The applicable analyses have typically run aground on the complexity of the full biomarker growth model, which is in principle a non-linear mixed-effects model of unknown structure, the fitting of which seems infeasible from realistically obtainable data.

**Methods:** It is known that to estimate Mean Duration of Recent Infection (MDRI) at a given value of the recent/non-recent -infection discrimination threshold, one may compress the full biomarker growth model into a relation capturing the probability of a recent test result as a function of time since infection. Noting that the time-derivative (gradient) of this curve (for a value of threshold – *h*) is identical to the formal likelihood relevant to Bayesian inference of the infection date, for a subject yielding an assay result * *h* * on the date of their first positive HIV test. This observation bypasses the need for fitting a complex detailed biomarker growth model. Using publicly available data from the CEPHIA collaboration, we calculated curves for a range of thresholds for the Sedia Lag assay and performed Bayesian inference of infection data, given a uniform prior implied by a last negative and first positive test.

**Results:** We demonstrate the generation of posteriors for infection date, for patients with various delays between their last negative and first positive HIV test, and a range of LAg assay results (ODn) hypothetically obtained on the date of the first positive result.

**Conclusion:** Depending on the last-negative / first-positive interval, there is a range of ODn values that yields posteriors significantly different from the uniform prior one would be left with based merely on interval censoring. Hence, a LAg ODn obtained on the date of, or soon after, diagnosis contains potentially significant information about infection dating. It seems worth analysing other assays with meaningful dynamic range, especially tests already routinely used in primary HIV diagnosis (for example chemiluminescent assays and reader/cartridge lateral flow tests which admit objective variable line intensity readings) which have a sufficient dynamic range that corresponds to a clinically meaningful range of times-since-infection that are worth distinguishing from each other.

## Introduction

There are many reasons to want to establish timelines of infection and disease progression in patients, across a variety of diseases. In the case of HIV, which is our current application, estimating the time of infectious exposure can assist in further investigations within studies of early pathogenesis, and also support difficult conversations during post test counselling, which touch on such sensitive matters as where and when one may have become infected, and whom one may have put at risk.

It is understood that such estimates, based on laboratory assays, will always be subject to significant uncertainty, most notably from inter subject variability of biological processes, but it seems important to formally assess what is and is not possible, given current biomarkers which provide some sort of calibratable clock for post-infection time. Numerous HIV infection biomarkers of this kind have been identified [1–4] though they are almost exclusively used for surveillance purposes. Despite their increasing use in clinical diagnosis [5], the use of infection-dating markers in individuals remains somewhat controversial – in part because of the lack of a systematic framework for the interpretation of such data.

One of the few well-known schemes for making some estimate of the stage or timing of infection, usually called the Fiebig staging system, provides for a number of categorical stages post infectious exposure, defined by various combinations of test positivity and test negativity of assays of varying detection sensitivity during early stages of viral replication and immune system response [6]. Most of the laboratory assays used to define these ‘Fiebig Stages’ are no longer routinely available, given the rapid evolution of the diagnostic industry, and so the original estimates of the location in time, relative to infectious exposure, of these stages are of limited practical application.

Recently, the underlying logic and statistical consistency of the concept of test-discrepancy have been more carefully elucidated by Grebe et al [6], leading to a generalisation of the Fiebig staging scheme. This generalisation provides for a flexible family of test-discordant states whose temporal meaning can be made precise with widely accessible data, which has also been curated for the benefit of potential users [7]. The notion of categorical infection ‘stages’ implies estimates of infection dates which are almost uniform probability/confidence distributions within boundaries set by test dates, with offsets implied by typical ‘diagnostic delays’ of the relevant tests [8]. In other words, while the ‘edges’ are not sharp, complete knowledge of last negative and first positive tests (ideally, but not necessarily, from different tests performed on the same date) is equivalent to the estimation of an interval in which infection is almost certainly located. Beyond the not entirely perfect boundaries of these intervals, there is little information, in such a scheme, about when, within that interval, infection is more likely to have occurred.

In order to go beyond these almost ‘pure interval’ estimates, the most obvious idea is to use the dynamic range of the biomarkers to hand, and to attempt to calibrate some informative dynamic range of values of these markers, to serve as some kind of approximate clock. While, as noted above, there is now increasing use of recent infection testing in clinical contexts, this is still largely based on categorical interpretation using some threshold [9,10], the choice (and not very clearly spelled out meaning) of which is informed from application to surveillance.

We adopt, according to Grebe et al 2016 [8] and Facente et al 2020 [11], the notion of ‘Estimated Date of Detectable Infection’ (EDDI), by which we mean the date at which the infection in a particular individual would first have been detectable, using a particular diagnostic algorithm being applied. (it has previously been used to specifically imply diagnosis by a sensitive viral load assay with a diagnostic threshold of one copy per ml of blood.) We will treat this, heuristically, as if it were the infection date, and thus avoid repeated cumbersome reference to the details of the formal procedure for incorporating the notion of assay-specific ‘diagnostic delays’ [8]. In the present context, these (heuristically bluntly interval censored) estimates are in any case merely used as Bayesian priors of infection date estimates, which precede knowledge of the continuously variable recency biomarker values.

In the present work, we demonstrate a simple approach, informed by precise formal statistical arguments, to generate continuously variable infection date estimates, when a sufficiently well characterised continuously variable infection marker is available from a test performed at or around the time of first objective evidence of infection.

Our investigation is substantially inspired by our work in using markers of infection timing to support surveillance – in particular incidence estimation. As we recycle not just the intuition about biomarkers developed for that purpose, but also analytical concepts, we briefly recap that:

- Incidence surveillance using markers of ‘recent infection’ is based on the heuristic that a large ‘prevalence’ of a categorically defined ‘recent infection’, among HIV positive respondents in a survey, is an indication of high incidence in a time window preceding the survey.
- This idea has been made precise by the work of Kassanjee et al 2012 [12], which defines the key concepts and analytical relationships that are required.
- A logically valid recent infection test can essentially be constructed from almost any continuous biomarker with a reasonable dynamic range post infection / initial detectability of infection.
- Whether a test for recent infection is usefully statistically informative of HIV incidence depends in considerable detail on many factors, including context, limitations of survey size, etc. but also, crucially, on the two key performance characteristics of the test (understood to be comprised of one or more assays, with rules for dichotomising the net result into *recent infection* versus *non-recent infection*):
  - The *Mean Duration of Recent Infection* (MDRI) is the mean time which individuals spend exhibiting the ‘recent’ range of the biomarker(s) employed in the test for recent infection. There is nominally a time cutoff, *T*, which is required to define the upper limit of ‘valid’ recent results – one of several fine points of statistical bookkeeping. The MDRI captures mainly the biology of early pathogenesis, and, in order for the recency test to be truly useful, should not have much variation between contexts.
  - The False Recent Rate (FRR) is the proportion of individuals who, despite being infected for longer than the time cut off *T*, nevertheless are classified as ‘recent’ by the laboratory procedures used to define that. At least for surveillance purposes, for this to be a tolerable feature of the test, the FRR should be very small (less than one percent), though it is expected to inevitably vary between contexts.

## Methods

In this analysis, we use publicly available data for the Sedia LAg assay published as part of Sempa et al 2019 [3]. From the 2424 samples in the CEPHIA Evaluation Panel, we used 928 unique samples, with a wide range of times since infection, mostly infected with HIV-1 subtype B (51%), C (28%), A1 (12%), and D (6%). The panel contains specimens from ART-suppressed individuals and the majority of subjects (68%) had sufficient clinical background data to produce Estimated Dates of Detectable Infection (EDDIs). These are infection time ‘point estimates’ accompanied by plausible intervals of first detectability, obtained by systematically interpreting diverse diagnostic testing histories according to the method previously described [8].

### Laboratory procedures

The CEPHIA Evaluation Panel was tested with the Sedia™ HIV-1 Limiting Antigen Avidity EIA (LAg) assay [13]. The LAg assay is microtitre-based with the solid phase of the microtitre plate coated with a multi-subtype recombinant HIV-1 antigen. This antigen is coated in a limiting concentration to prevent crosslinking of antibody binding, making it easier to remove weakly-bound antibody. Specimen dilutions are incubated for 60 minutes and then a disassociation buffer is added for 15 minutes to remove any weakly-bound antibody. A goat anti-human, horseradish peroxidase (HRP)-conjugated IgG is added and this binds to any remaining IgG; a tetramethylbenzidine substrate is added and a colour is generated which is proportionate to the amount of HRP. An optical density (OD) is measured for each sample and then normalized by the use of a calibrator specimen. On each plate, the calibrator is tested in triplicate, with the median of the three ODs used to normalize specimen readings, producing normalized optical density (ODn) measurements. The Sedia LAg testing procedure requires that specimens producing an initial ‘screening’ OD of ≤2.0 be subjected to triplicate confirmatory testing.

### Statistical analysis

The key point of the present analysis is to demonstrate

- how it is possible to perform a fully-fledged Bayesian inference of infection date, for subjects who are newly diagnosed at a known time after a last negative diagnostic test result, if,
- on or near the date of the first positive diagnosis, there is also a result from a well characterised and calibrated continuous biomarker assay with a ‘sufficient’ dynamic range.

The fundamental requirements for performing a Bayesian analysis are:

- A ‘**prior’ distribution of the target variable**, i.e. a distribution governing what is known about the target variable, even before the particular context-specific crucial experimental result is known.
- A fully specified ‘**likelihood’ function**, which gives the probability (or probability density, for continuous variables) of the biomarker, given a hypothetical value of the target variable (for example, probability density of obtaining a LAg ODn value *h*, given that the subject has been infected for time *t*).

In the present case, as we are interested in someone about whom it is initially known that they had a most recent negative test at some time, and a first positive diagnosis some time thereafter, the **prior distribution** on infection time is (almost) just the interval between the last negative and first positive test. To be more precise, there is an offset at each end (not necessarily the same amount – a subtle point explained elsewhere [8]) to account for ‘diagnostic delay’ – i.e. typical, delay, given the applicable diagnostic algorithm, between *infection* (infectious exposure) and algorithm-specific *detectability of infection*.

We are not aware that the analysis which follows, to calculate the **likelihood**, has been previously presented, though there have been numerous efforts to summarise biomarker progression into what would traditionally be formally called (non-linear, probably) ‘mixed effects models’. This is a technical way of saying that:

- The biomarker follows some potentially complex function over time
- The ultimate biomarker values attained by a laboratory assay would additionally have some kind of measurement noise.
- The biomarker ‘trajectory’ is different for different people
- Individual ‘trajectories’ can be simulated via a specific functional form, with parameters (scale and shape parameters, asymptotes, etc.) drawn from certain distributions.

This is indeed a very broad family of behaviours, and it is logically sufficient to describe an arbitrarily complex biomarker. It also leads to severe (we suspect, unsurmountable) challenges to those wishing to fit the parameters of an appropriate functional form to available data. Certainly, if a fully specified model of biomarker growth were somehow available, it would provide a fully specified likelihood function, in the sense that this model would determine the probability (density) of attaining a biomarker result *h*, at some time *t* post infection. The fully specified model would also implicitly specify many other things, including correlations between measurements from an individual via samples taken at particular (different) times. We show below that because we require just likelihood of obtaining a single observation, and are not currently interested in a larger complex set of statistical metrics, we do not need the full biomarker evolution model!

In order to estimate the MDRI (see above) at a given value of the recent/non-recent -infection discrimination threshold, one can *in principle*:

- obtain a fully specified biomarker regression model (though, as noted above, this is no small task, and may actually not be possible in practice)
- perform the relevant integrations over all possible realisations of the biomarker value at a time *t* post infection, and hence
- derive the mean time which individuals spend displaying the ‘recent’ range of values.

Given the keen interest in estimates of MDRI for various biomarkers, for the purpose of estimating HIV incidence, there has been some investment of thought in how to circumvent reliance on a detailed, difficult-or-impossible-to-obtain, biomarker progression model. It has been shown Kassanjee et al 2017 [14] that one may robustly estimate an MDRI (for a given assay and specified value of the recent/non-recent threshold, *h*) by empirically compressing the full biomarker growth model into a relation capturing just the probability of obtaining a *recent* (vs *non-recent*) test result, as a function of time since infection. This function has previously been denoted *P*_*r*_(*t*|*h*), and it can be fitted, without much fanfare, to data of the kind that is in fact available from studies which have intensively followed up individuals in the weeks and months post diagnosis.

Typically, the raw data from such studies provides numerous observations (recency test results) of people who have a last negative diagnostic test not too long before a first positive test, making the time of infection known with reasonable precision (typically locating the time of infection within a window of a several weeks). The recency test results are prototypically based on continuously variable values of some underlying biomarker, which is reduced to a recent/non-recent verdict by use of a threshold.

For each threshold value, *h*, of interest, one needs to perform the dichotomisation of the recency marker data, and then perform some sort of bionomial regression which leads to a (in practice, parametrized) function *P*_*r*_(*t*|*h*). Mathematically, this is just a function of two variables - *h* and *t*. To produce this function with a reasonable level of ‘resolution’ we need to merely pick a sufficiently narrowly spaced set of values of *h*, and perform the fitting procedure for each of these recent/non recent thresholds. Now we note that:

- a *likelihood density* is nothing more than the *derivative* of a *cumulative likelihood*
- the function *P*_*r*_(*t*|*h*), read as a function of *h*, is the probability of obtaining values below *h*, at time *t*.
- so the *likelihood density* of obtaining biomarker values near *h*, at time *t* post infection, is given by 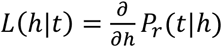

Given the construction of *P*_*r*_(*t*|*h*), the variation in the *t* direction can be obtained analytically from the parametrisation of the fitting procedure. However, the required derivative is with respect to *h*, and variation in the *h* direction is not analytically available, since the function is fitted separately for each value of *h*. This is merely a numerical problem, and there are in practice many ways to obtain these derivatives with sufficient numerical smoothness in order to produce plots of the likelihood, and posteriors.

We obtained a family of *P*_*r*_ (*t*|*h*) functions using a binomial regression with a logit link function and up to cubic terms in time. We stored the fitted parameters for various values of LAG ODn threshold, *h*, from 0.1 to 4, in steps of 0.01. We want to evaluate likelihood densities some combination of *h*_*0*_ (the observed ODn which forms the basis of an infection time estimate) and *t* (which takes various values in the range of plausible times-since-infection). To do this, we

- look up the precomputed values *P*_*r*_(*t*|*h*) at the target value of *t*, for the nearest four stored values of *h* (nearest *h*_0_).
- fit *P*_*r*_(*t*|*h*) as a cubic spline **in the *h* direction**, and
- evaluate the derivative, with respect to *h*, of this fitted spline, at *h*_0_.

This provides fast and smooth evaluation of the likelihood densities which are required to perform the infection time inference.

In order to demonstrate the application to infection time inference, we considered hypothetical newly diagnosed individuals with a range of intervals between a last negative and first positive diagnostic test. In practice, there is a delay between infectious exposure and detectability of infection, but this is a point which we do not consider in detail here – it will merely require adding (usually slightly different) offsets on each end of the interval over which the bayesian ‘prior’ for infection time is taken to be uniform. Then, adding the recency marker value as the critical additional information, and using the likelihood density constructed as outlined, we obtain posteriors, which in this case (uniform priors) are simply the correctly normalised likelihood densities.

Statistical analysis was done using R, version 4.0.2 (R Foundation for Statistical Computing, Vienna, Austria)

## Results

Figure 1 shows, as a scatterplot, the values of ‘time since EDDI’ and Sedia LAg Avidity normalised optical density (ODn) for the specimens in the CEPHIA evaluation panel data set which were used for the present analysis. The clustering in the time direction not surprising, and arises from the estimation of EDDI as the midpoint of the (often whole months) interval between last negative test and first positive diagnostic test.

**Figure 1:**
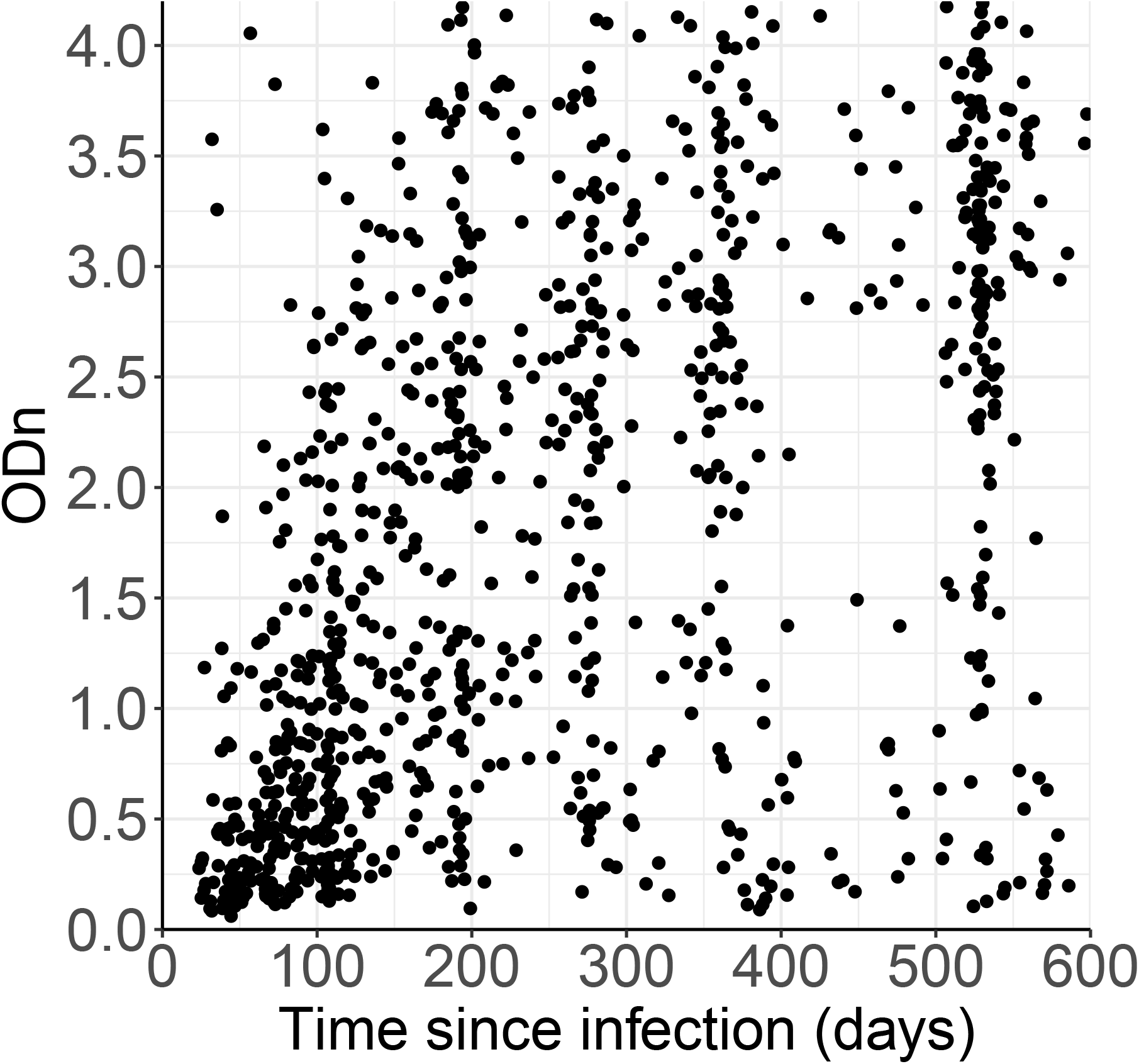
Distribution of ODn vs days since EDDI of CEPHIA evaluation panel specimens from subejcts who were HIV positive, ART-naïve and non-elite controllers.

Figure 2 shows *P*_*r*_(*t*) curves for the LAg assay, out to 600 days, for a range of threshold to define recent infection. At lower ‘recency’ thresholds, the *P*_*r*_(*t*) curves approach zero convincingly, while for higher ODn values (roughly ODn >2) they clearly do not approach zero.

**Figure 2:**
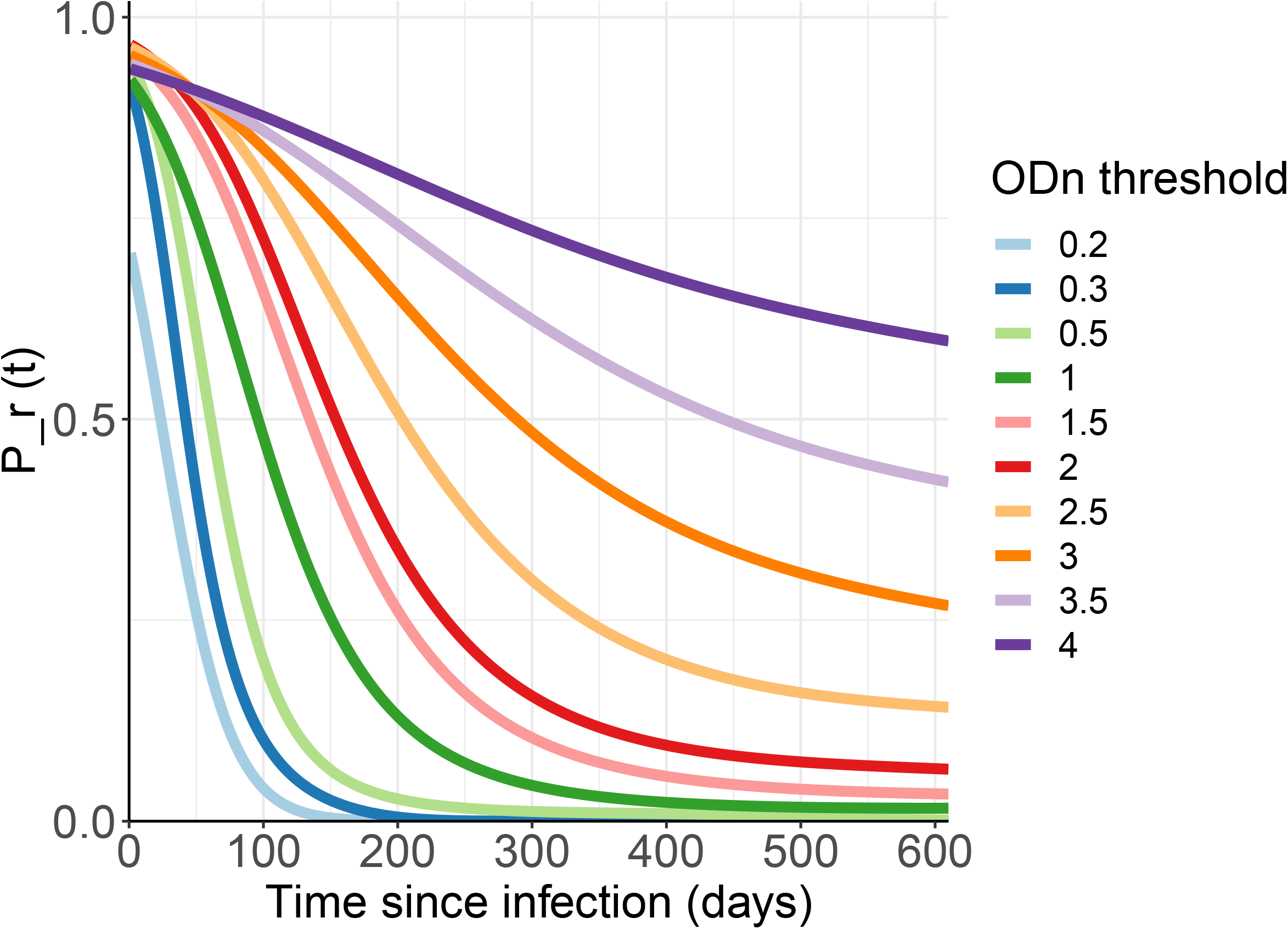
Probability of exhibiting the recent marker as a function of time since detectable infection.

Using the logic described in the methods section, we transformed the *P*_*r*_(*t*) curves into likelihoods. Figure 3 captures these likelihoods through the lens of percentile ‘growth curves’ for Sedia LAg ODn, which can be read much as one reads baby growth curves for height and weight, for example. Given a value of time since infection (X axis), a vertical slice through the percentile curves indicates what percentage of subjects’ ODn values lie below the Y values of the contours as they cross the chosen X value. These growth curves are one obvious way one might interpret/present ODn values obtained at the date of first diagnosis. However, this is still a fundamentally frequentist presentation, telling us the probabilities of seeing various values of ODn, given a time since infection, without telling us the probabilities of hypothetical values of time since infection, given an ODn. Hence, we find the Bayesian posterior approach more appealing.

**Figure 3:**
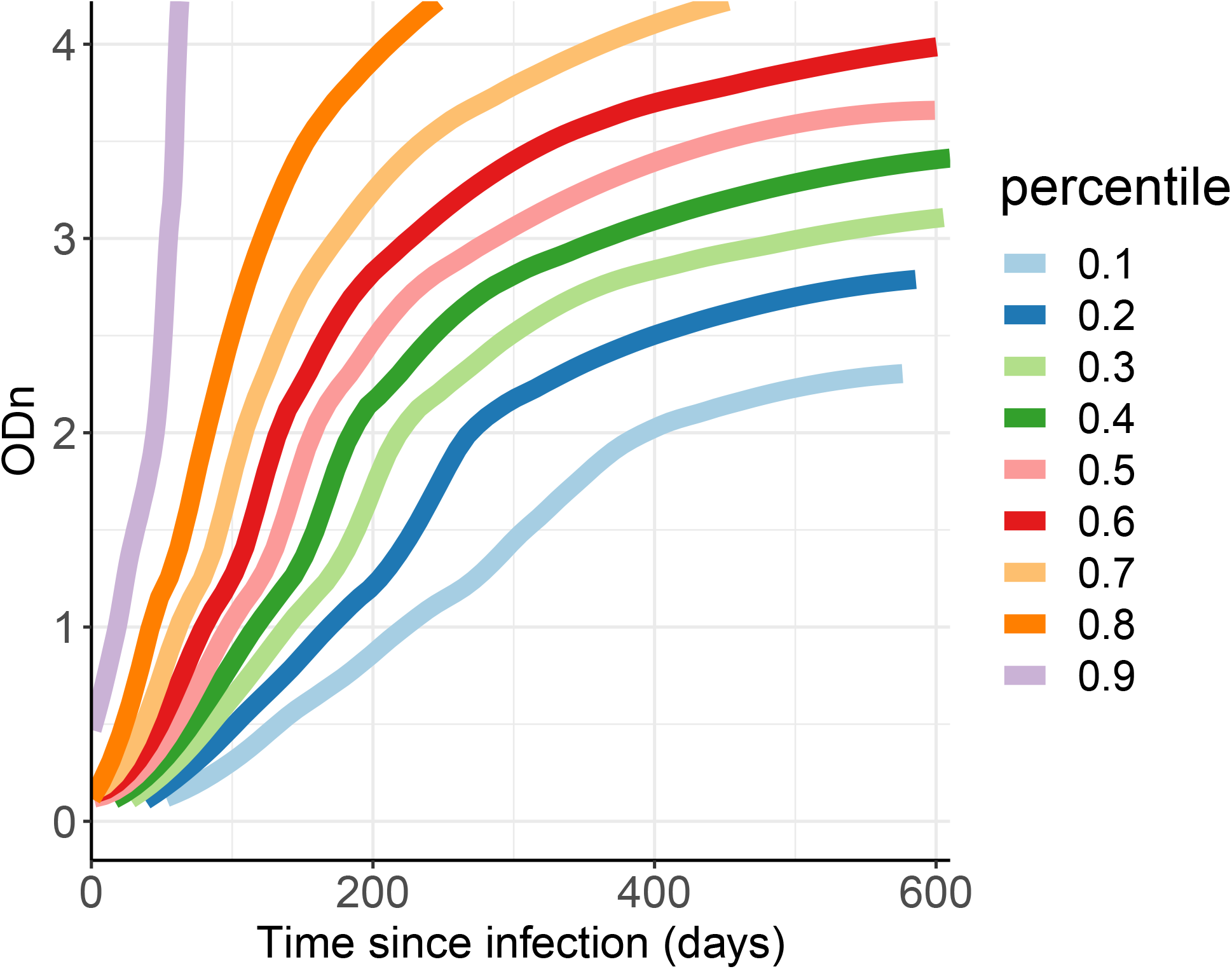
Percentile curves generated from probability of exhibiting the recent marker as a function of time since detectable infection--*P*_*r*_(*t*).

In Figure 4, we present the posterior density for a selection of values of ODn (0.2, 0.3, 0.5, 1.5 and 4) obtained on the day of first positive test, for different inter-test intervals (50, 100, 300 and 600 days). In each case, the EDDI point estimate is the midpoint of the inter-test interval. As expected, not all values of ODn are equally informative, and the value of the ODn data varies significantly with the inter-test interval. Note that in our case, where the prior distribution is always uniform, the shape of the posteriors is always the same as the likelihood itself, viewed/sliced as a function of time since infection. The families of curves look slightly different between the panels of figure 4 because in each case we normalise the posterior sensibly to provide a cumulative probability of 1 over the inter-test interval. Figure 5 applies to the same scenario as panel D in Figure 4, but reveals a little more detail through the inclusion of a few additional values of ODn.

**Figure 4:**
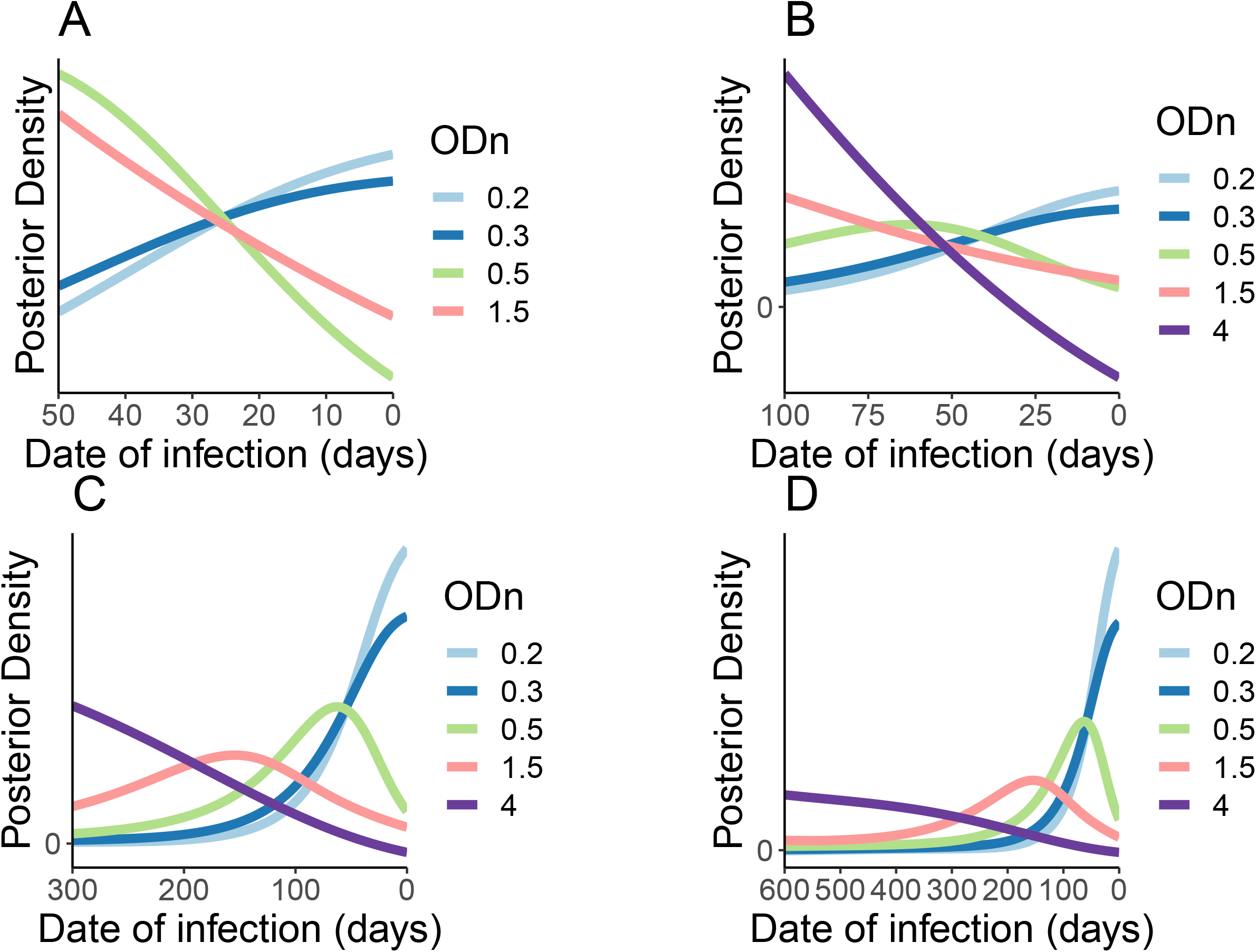
Posterior distribution of various ODn values and inter-test intervals of last negative and first positive HIV test, i.e. *L*(*h*|*t*). **Legend**: **A**—inter-test interval of 50 days; **B**— inter-test interval of 100 days; **C**—inter-test interval of 300 days; **D**— inter-test interval of 600 days.

**Figure 5:**
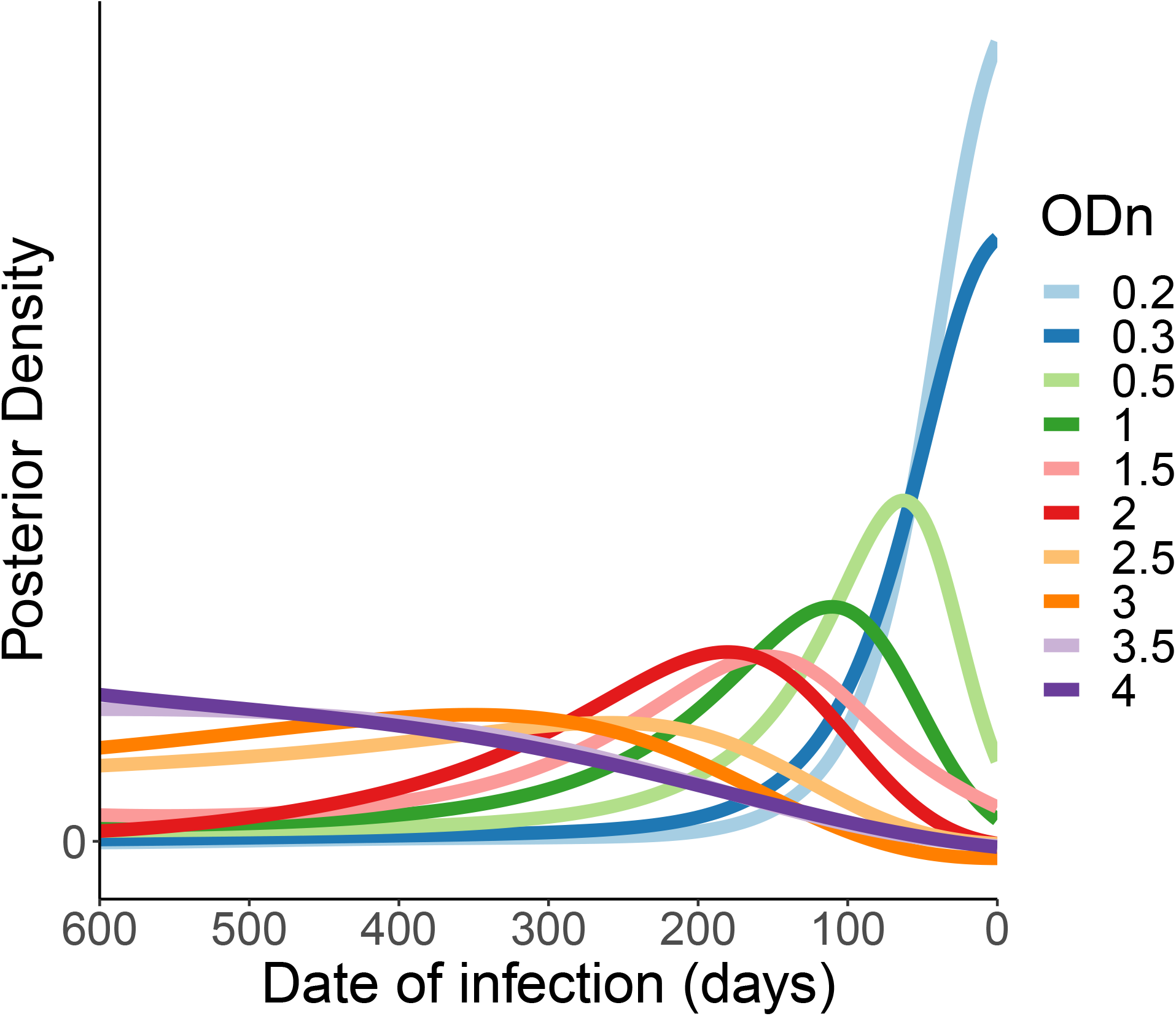
Posterior distribution of various ODn values for an inter-test interval of 600 days.

Note that not all posteriors have a well-defined mode inside the sensible range of values. This merely captures that for some situations there is no internal ‘most likely’ region for the infection date – it is merely increasingly likely to lie ever further to one side, within the limits sets by the diagnostic tests.

## Discussion

Our ‘efficient’ derivation of the likelihood density, which is the crux of infection date estimation procedure, is nothing more than a usually neglected interpretation of constructs which have long been routinely used to estimate MDRI for putative recent infection tests. It does not rely on any special or controversial assumptions. Whether there is sufficient data to estimate this likelihood density without significant parametric artifacts will be a question that must be investigated for any particular biomarker intending to be used in infection timing estimates of the kind we are proposing here. For previously described markers such as the Sedia LAg avidity assay, and numerous others which have been well studied by both an original developer and an independent laboratory [1–4], it seems clear that sufficient data is available to use the proposed method outlined here for estimating infection dates.

Given a particular biomarker, we believe that the fundamental analysis developed here presents the most efficient/informative extraction of infection date posteriors that one can hope for. How useful these estimates are, in practice, will depend on a number of details which we do not claim to be investigating at the present time. In particular:

- We propose that there be some qualitative investigation into how either the ‘percentile curves’ or the ‘posterior infection date’ estimates can be used in clinical settings, to ascertain how patients and healthcare providers understand and value this information, and hence to propose meaningful guidelines for clinicians to speak about them.
- Details of biological studies and early intervention trials will determine how significantly more useful the fully developed posterior estimates are, compared to the priors obtained by the post Fiebig interval censoring of Grebe et al 2019 [8].

Given that the statistical ‘heavy lifting’ is done once, up front, when fitting *P*_*r*_(*t*|*h*) for various values of *h*, the computational effort involved in generating these estimates for individual patients is very modest. For a serious study of utility in a clinical context, we have developed an Rshiny app, which requests

- the last-negative / first positive test interval,
- (optionally) the specification of the diagnostic tests, or their mean ‘diagnostic delays’, and
- the specification, and value, of the recency marker obtained at or near the time of first diagnosis

and, from this information, generates the infection date posterior. To access the R Shiny app please follow this link: https://jbsempa.shinyapps.io/bm_posteriors_pctcurves/. Further, we are in the process of expanding it to include other recency like Maxim LAg, Abort Architect and some annotation like specific percentile ranges.

Despite the hesitancy in some quarters about using recent infection tests for individual level rather than population level testing [15], we note that it is increasingly being done, and will continue to be done, whether or not there is consensus on its meaning and appropriate use. We would argue that the formalisation of Bayesian infection date estimates, which we have developed here, is a crucial component of a rational discussion about such applications.

Despite the hesitancy in some quarters about using recent infection tests for individual level rather than population level testing [15], we note that it is increasingly being done, and will continue to be done, whether or not there is consensus on its meaning and appropriate use. We would argue that the formalisation of Bayesian infection date estimates, which we have developed here, is a crucial component of a rational discussion about such applications.

## Data Availability

All the data used in this manuscript was already published in a previous article PMID: 31348809.

https://doi.org/10.1371/journal.pone.0220345.s008

## Acknowledgement

This work is based on research supported by the (South African) Department of Science and Innovation and the National Research Foundation. Any opinion, finding, and conclusion or recommendation expressed in this material is that of the authors and the NRF does not accept any liability in this regard.

## References

1 Kassanjee R, Pilcher CD, Keating SM, Facente SN, McKinney E, Price MA, et al. Independent assessment of candidate HIV incidence assays on specimens in the CEPHIA repository. Aids 2014; 28:2439–2449.

2 Kassanjee R, Pilcher CD, Busch MP, Murphy G, Facente SN, Keating SM, et al. Viral load criteria and threshold optimization to improve HIV incidence assay characteristics. Aids 2016; 30:2361–2371.

3 Sempa JB, Welte A, Busch MP, Hall J, Hampton D, Facente SN, et al. Performance comparison of the Maxim and Sedia Limiting Antigen Avidity assays for HIV incidence surveillance. PLoS One 2019; 14:e0220345.

4 Keating SM, Kassanjee R, Lebedeva M, Facente SN, MacArthur JC, Grebe E, et al. Performance of the Bio-Rad Geenius HIV1/2 Supplemental Assay in Detecting “Recent” HIV Infection and Calculating Population Incidence. J Acquir Immune Defic Syndr 2016; 73:581–588.

5 Hassan J, Moran J, Murphy G, Mason O, Connell J, De Gascun C. Discrimination between recent and non-recent HIV infections using routine diagnostic serological assays. Med Microbiol Immunol 2019; 0:0.

6 Fiebig EW, Wright DJ, Rawal BD, Garrett PE, Schumacher RT, Peddada L, et al. Dynamics of HIV viremia and antibody seroconversion in plasma donors: implications for diagnosis and staging of primary HIV infection. AIDS 2003; 17:1871–9.

7 Ananworanich J, Fletcher JLK, Pinyakorn S, van Griensven F, Vandergeeten C, Schuetz A, et al. A novel acute HIV infection staging system based on 4th generation immunoassay. Retrovirology 2013; 10:56.

8 Grebe E, Facente SN, Bingham J, Pilcher CD, Powrie A, Gerber J, et al. Interpreting HIV diagnostic histories into infection time estimates: analytical framework and online tool. BMC Infect Dis 2019; 19:894.

9 Robinson E, Moran J, O’Donnell K, Hassan J, Tuite H, Ennis O, et al. Integration of a recent infection testing algorithm into HIV surveillance in Ireland: improving HIV knowledge to target prevention. Epidemiol Infect 2019; 147. doi:10.1017/S0950268819000244

10 Rice BD, de Wit M, Welty S, Risher K, Cowan FM, Murphy G, et al. Can HIV recent infection surveillance help us better understand where primary prevention efforts should be targeted? Results of three pilots integrating a recent infection testing algorithm into routine programme activities in Kenya and Zimbabwe. J Int AIDS Soc 2020; 23 Suppl 3:e25513.

11 Facente SN, Grebe E, Pilcher CD, Busch MP, Murphy G, Welte A. Estimated dates of detectable infection (EDDIs) as an improvement upon Fiebig staging for HIV infection dating. Epidemiol Infect 2020; 148:e53.

12 Kassanjee R, McWalter TA, Bärnighausen T, Welte A. A new general biomarker-based incidence estimator. Epidemiology 2012; 23:721–8.

13 Sedia Biosciences Corporation. Sedia™ HIV-1 LAg-Avidity EIA: single well avidity enzyme immunoassay for detection of recent HIV-1 infection, Cat. No. 1002. ; 2016. http://www.sediabio.com/LiteratureRetrieve.aspx?ID=134692

14 Kassanjee R, De Angelis D, Farah M, Hanson D, Labuschagne JPL, Laeyendecker O, et al. Cross-Sectional HIV Incidence Surveillance: A Benchmarking of Approaches for Estimating the “Mean Duration of Recent Infection”. Stat Commun Infect Dis 2017; 9. doi:10.1515/scid-2016-0002.

15 The Foundation for AIDS Research. New HIV Testing Strategies in PEPFAR COP19: Rollout and Human Rights Concerns. Washington DC.: ; 2019. https://www.amfar.org/uploadedFiles/_amfarorg/Articles/On_The_Hill/2019/COP19.pdf (Accessed 27 Mar2019).

